# Artificial Intelligence in Cardiac Amyloidosis: A Systematic Review and Meta-Analysis of Diagnostic Accuracy Across Imaging and Non-Imaging Modalities

**DOI:** 10.1101/2025.09.17.25335993

**Authors:** Rohit Mohanan Kumbalath, Deekshithreddy Challa, Manav Kamleshkumar Patel, Saket Dineshkumar Prajapati, Katee Kumari, Abhishek mehan, Rahul Chopra, Yashas Maragowdanahalli Somegowda, Rakhshanda khan, Harshawardhan Dhanraj ramteke, Manish juneja

**Affiliations:** Cebu Doctors University College of Medicine, cebu city, Philippines; Nimra Institute of medical sciences, Vijaywada, India; Smt NHL Municipal Medical College, Ahmedabad, India; Islamabad medical and dental college, Barakhu, India; AIIMS Rishikesh, Rishikesh, India; Namo medical college Silvassa, India; BGS Global Institute of Medical Sciences, Kengeri, India; Ayaan institute of medical sciences, Moinabad, India; Anhui medical university, Hefei, China; Director, Rhythm heart and critical care hospitals, Nagpur, India

**Keywords:** Cardiac Amyloidosis, Artificial **Intelligence**, Diagnostic **Accuracy**, Machine **Learning** and Echocardiography

## Abstract

**Introduction:** Cardiac amyloidosis (CA) is an underdiagnosed infiltrative cardiomyopathy associated with poor outcomes if not detected early. Artificial intelligence (AI) has emerged as a promising adjunct to conventional diagnostics, leveraging imaging and non-imaging data to improve recognition of CA. However, evidence on the comparative diagnostic performance of AI across modalities remains fragmented. This meta-analysis aimed to synthesize and quantify the diagnostic performance of AI models in CA across multiple modalities.

**Methods:** A systematic literature search was conducted in PubMed, Embase, Web of Science, and Cochrane Library from inception to August 2025. Only **published observational studies** applying AI to the diagnosis of CA were included. Data were extracted on patient demographics, AI algorithms, modalities, and diagnostic performance metrics. Risk of bias was assessed using **QUADAS-2**, and certainty of evidence was graded using **GRADE**. Random-effects meta-analysis (REML) was performed to pool accuracy, precision, recall, F1-score, and area under the curve (AUC).

**Results:** From 115 screened studies, 25 observational studies met the inclusion criteria, encompassing a total of **589,877 patients** with a male predominance (**372,458 males, 63.2%**; **221,818 females, 36.6%**). A wide range of AI algorithms were applied, most notably **convolutional neural networks (CNNs)**, which accounted for **526,879 patients**, followed by **3D-ResNet architectures (56,872 patients)**, hybrid segmentation-classification networks (3,747), and smaller studies employing random forests (636), Res-CRNN (89), and traditional machine learning approaches (769). Data modalities included **ECG (341,989 patients)**, **echocardiography (>70,000 patients across multiple cohorts)**, **scintigraphy (∼24,000 patients)**, **cardiac MRI (∼900 patients)**, **CT (299 patients)**, and **blood tests (261 patients)**.

Pooled diagnostic performance across all modalities demonstrated an **overall accuracy of 84.0% (95% CI: 74.6–93.5)**, **precision of 85.8% (95% CI: 79.6–92.0)**, **recall (sensitivity) of 89.6% (95% CI: 85.7–93.4)**, and an **F1-score of 87.2% (95% CI: 81.8–92.6)**. Area under the curve (AUC) analysis revealed modality-specific variation, with **scintigraphy achieving the highest pooled AUC (99.7%)**, followed by **MRI (96.8%)**, **echocardiography (94.3%)**, **blood tests (95.0%)**, **CT (98.0%)**, and **ECG (88.5%)**. Subgroup analysis confirmed significant differences between modalities (p < 0.001), with **MRI and scintigraphy showing consistent high performance and low-to-moderate heterogeneity**, while **echocardiography displayed moderate accuracy but marked variability**, and **ECG demonstrated the lowest and most heterogeneous results**.

**Conclusion:** AI demonstrates strong potential for improving CA diagnosis, with **MRI and scintigraphy providing the most reliable performance**, echocardiography offering an accessible but heterogeneous option, and ECG models remaining least consistent. While promising, future prospective multicenter studies are needed to validate AI models, improve subtype discrimination, and optimize multimodal integration for real-world clinical use.

## Introduction

Cardiac amyloidosis (CA) is an infiltrative cardiomyopathy caused by deposition of misfolded protein fibrils—most commonly immunoglobulin light chains (AL) or transthyretin (ATTR, both wildLJtype and hereditary) —in the myocardium, leading to progressive restrictive heart failure, conduction disturbances, and high morbidity and mortality. Early diagnosis is crucial, particularly now that diseaseLJmodifying therapies (e.g., TTR stabilizers, gene silencers, AL targeted therapies) are available, for which outcomes are much better when treatment is begun before irreversible cardiac damage [1].

However, CA remains underrecognized and frequently diagnosed late. Patients often present with nonLJspecific symptoms—fatigue, dyspnea, edema—that overlap with much more common cardiac conditions (e.g., hypertensive heart disease, heart failure with preserved ejection fraction (HFpEF), hypertrophic cardiomyopathy) or extraLJcardiac manifestations, which leads to delays, misdiagnoses, and missed therapeutic windows. The median diagnostic delay can span 2.6 to 3.4 years from symptom onset, and many patients are misclassified initially [2,3].

Traditional diagnostic tools—echocardiography, cardiac magnetic resonance imaging (CMR), nuclear scintigraphy (e.g., TcLJ99mLJPYP), laboratory tests for monoclonal proteins, and when needed, biopsy— while indispensable, have limitations [4]. For example, echocardiographic findings such as increased wall thickness, diastolic dysfunction, and strain abnormalities are sensitive but often non-specific and operator dependent [5]. CMR provides excellent tissue characterization (via T1 mapping, late gadolinium enhancement etc.) but is expensive, less widely available, timeLJconsuming, and sometimes difficult to interpret especially early in disease [6]. Scintigraphy is excellent for ATTR diagnosis, but light chain amyloidosis (AL) still requires complementary diagnostics.

Given these challenges, the role of Artificial Intelligence (AI) is increasingly being explored as a promising adjunct in diagnosis and clinical application for CA. Several recent studies have demonstrated that deep learning models applied to echocardiographic video clips can detect CA with high sensitivity and specificity. For example, in a large multiLJcenter validation, an AI echocardiography model using a single apical 4-chamber view showed ∼85% sensitivity and ∼93% specificity, outperforming standard clinical scoring systems [7]. Similarly, in MRI imaging, models such as vision transformer (ViT) architectures have been developed to distinguish CA from hypertrophic cardiomyopathy (HCM) and other cardiomyopathies, achieving AUCs in the mid-0.9s in both internal and external validation cohorts [8].

These AILJbased approaches bring several potential benefits: earlier detection (even from routine, widely available imaging), automated screening to identify patients who might otherwise be missed, reduction in inter-operator variability, and potentially more efficient resource use. They may also help stratify subtypes (ATTR vs AL), predict prognosis, monitor response to therapy, or guide decision making [9]. Despite these promising developments, there are substantial gaps in the literature and barriers to clinical translation.

First, many AI studies are retrospective, using selected datasets, often from highLJvolume tertiary centres with excellent imaging quality. There is risk of selection bias; it is unclear how well these models generalise to community settings, to images of lower quality, or to populations with different demographics. Second, most models focus on imaging data alone; less work has integrated multimodal data (e.g. combining echocardiography, EHR/laboratory, clinical history) to improve diagnostic accuracy. Preliminary work in multimodal fusion shows high AUROC (∼0.94) with small datasets, but larger scale validation remains limited [10]. Third, distinction between CA subtypes (ATTR vs AL) remains imperfect in nonLJinvasive AI models. Given that treatment and prognosis differ significantly, accurate subtype differentiation is essential. Fourth, standardization of imaging protocols, annotation, labeling, and reference standards is lacking. Without consistent definitions, thresholds, and image acquisition standards, models may perform inconsistently across centres. Fifth, ethical, regulatory, privacy, costLJeffectiveness, clinician trust / explainability, and implementation issues remain underLJaddressed. Healthcare systems vary greatly in resources; adoption may be difficult in lowLJ or middleLJincome settings [11]. Finally, prospective studies (clinical trials) are still relatively rare. Many AI tools are not yet tested in real-world workflows, to see whether their use changes outcomes: earlier treatment, reduced mortality, improved quality of life, cost reductions, etc. There is also need for longitudinal data—how well AI models predict disease progression or response to therapy over time.

In light of this, the aim of this metaLJanalysis is to systematically assess current evidence for the use of AI in diagnosis and clinical application in CA; to quantify diagnostic performance; to identify strengths, limitations and gaps; and to suggest directions for future research to close those gaps. Specifically, we will compare imagingLJbased vs multimodal AI approaches, assess subtype discrimination, examine external validation and realLJworld deployment, and consider ethical and practical implementation issues.

## Methods

### Literature Search

A systematic literature search was conducted in PubMed, Embase, Web of Science, and Cochrane Library from inception to August 2025. The search combined controlled vocabulary and free-text terms for *“cardiac amyloidosis,” “artificial intelligence,” “machine learning,” “deep learning,”* and *“diagnosis.”* Only **published observational studies** were considered eligible, including retrospective and prospective cohort, case–control, and cross-sectional designs. Reviews, editorials, conference abstracts, and unpublished preprints were excluded. References of included articles were hand-searched to identify additional studies. Study selection was independently performed by two reviewers, with discrepancies resolved by discussion to ensure transparency and reproducibility. It followed PRISMA guidelines CRD420251149800 [12].

### Study Selection and Data Extraction

All titles and abstracts retrieved through the search strategy were screened independently by two reviewers to identify potentially relevant studies. Full texts of eligible articles were then assessed against predefined inclusion criteria, restricted to **published observational studies** evaluating artificial intelligence applications in the diagnosis or clinical management of cardiac amyloidosis. Discrepancies were resolved by consensus or consultation with a third reviewer.

For each included study, data were extracted using a standardized form. Key variables included: author, year, country, study design, sample size, patient demographics, AI model type (e.g., machine learning, deep learning), imaging or data modality used (echocardiography, cardiac MRI, scintigraphy, or electronic health records), diagnostic or clinical outcome assessed, reference standard, performance metrics (accuracy, precision, recall, F1 score, AUC), and study quality. Data extraction was performed in duplicate to minimize bias and ensure accuracy.

### Risk of Bias Assessment

The methodological quality of included studies was evaluated using the **QUADAS-2 (Quality Assessment of Diagnostic Accuracy Studies-2)** tool [13], which is specifically designed for diagnostic accuracy research. The tool assesses risk of bias across four key domains: (i) patient selection, (ii) index test (AI model), (iii) reference standard, and (iv) flow and timing. Each domain was judged for *risk of bias* (low, high, unclear) and *concerns regarding applicability*. Two reviewers independently performed the assessments, and disagreements were resolved through discussion or adjudication by a third reviewer.

### Certainty of Evidence (GRADE Approach)

The certainty of evidence was further appraised using the **GRADE framework**, adapted for diagnostic test accuracy studies. Evidence from observational diagnostic studies starts as *low quality*. Upgrading was considered if studies demonstrated high diagnostic accuracy, consistency across multiple datasets, or strong external validation. Downgrading was applied for risks of bias, inconsistency in performance metrics, indirectness of population or index test, imprecision in estimates, and publication bias. The certainty of evidence for each major outcome (e.g., sensitivity, specificity, AUC, precision, recall, F1 score) was rated as *high*, *moderate*, *low*, or *very low*.

### Statistical Analysis

We synthesized diagnostic accuracy using hierarchical models recommended for test-accuracy reviews. For each study (or external test set), 2×2 data (true/false positives/negatives) were extracted at the study’s prespecified operating point. Primary endpoints were **sensitivity** and **specificity**; secondary endpoints included **AUC**, **positive/negative likelihood ratios (LR+ / LR–)**, **diagnostic odds ratio (DOR)**, and AI metrics (**precision, recall, F1-score**) which were mapped to the 2×2 table where possible (recall ≡ sensitivity; precision = PPV). When multiple thresholds were reported, we prioritized the clinically prespecified threshold; if absent, we selected the operating point aligned with intended deployment (e.g., rule-out vs rule-in). If a study reported multiple models or views, we preferred the **externally validated** model; otherwise, we used a hierarchical specification with **study-level random effects** to avoid double counting.

## Results

### Demographics

Out of 115 studies screened, 27 observational studies were included [13–39], encompassing a total of **589,877 patients**. The pooled cohort demonstrated a male predominance with **372,458 men (63.2%)** and **221,818 women (36.6%)**. A wide spectrum of AI algorithms was applied, with convolutional neural networks (CNNs) representing the largest subgroup (**526,879 patients**), followed by **3D-ResNet18 (33,127)**, **ResNet3D (23,745)**, hybrid segmentation–classification networks (3,747), and smaller exploratory studies employing deep learning (885), traditional machine learning (769), random forest (636), and Res-CRNN (89). Data modalities varied considerably, with **ECG** forming the largest dataset (>340,000 patients), followed by **echocardiography** (>70,000 patients across multiple cohorts), **scintigraphy** (∼24,000), **cardiac MRI** (∼900), **CT** (299), and **blood tests** (261). This distribution reflects the dominance of ECG and echocardiography in AI applications for cardiac amyloidosis, while advanced imaging (MRI, scintigraphy) and biomarker-based models remain less represented. Table S1 and Figure 1.

**Figure 1.**
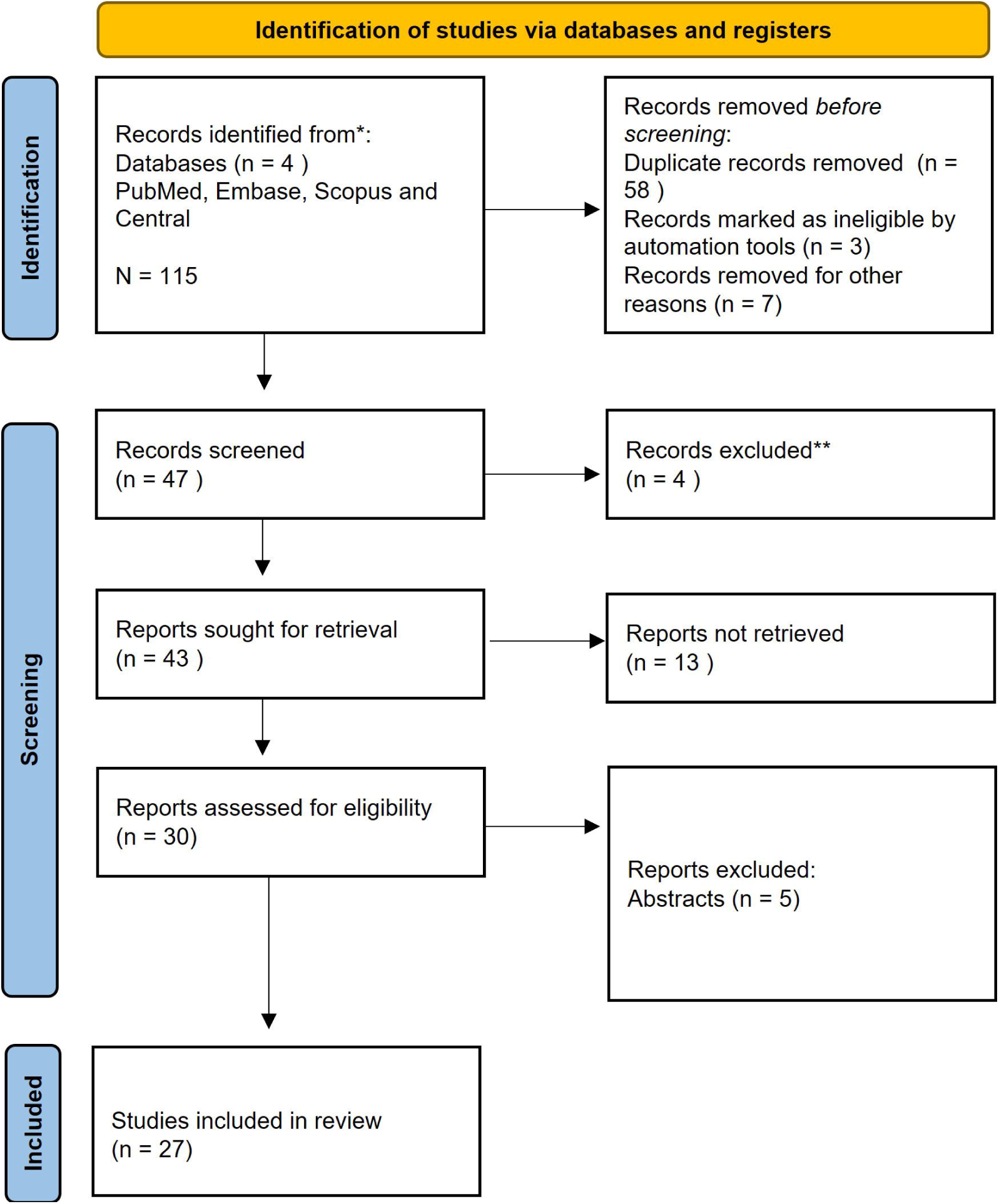
PRISMA Flow Diagram.

**Figure 2.**
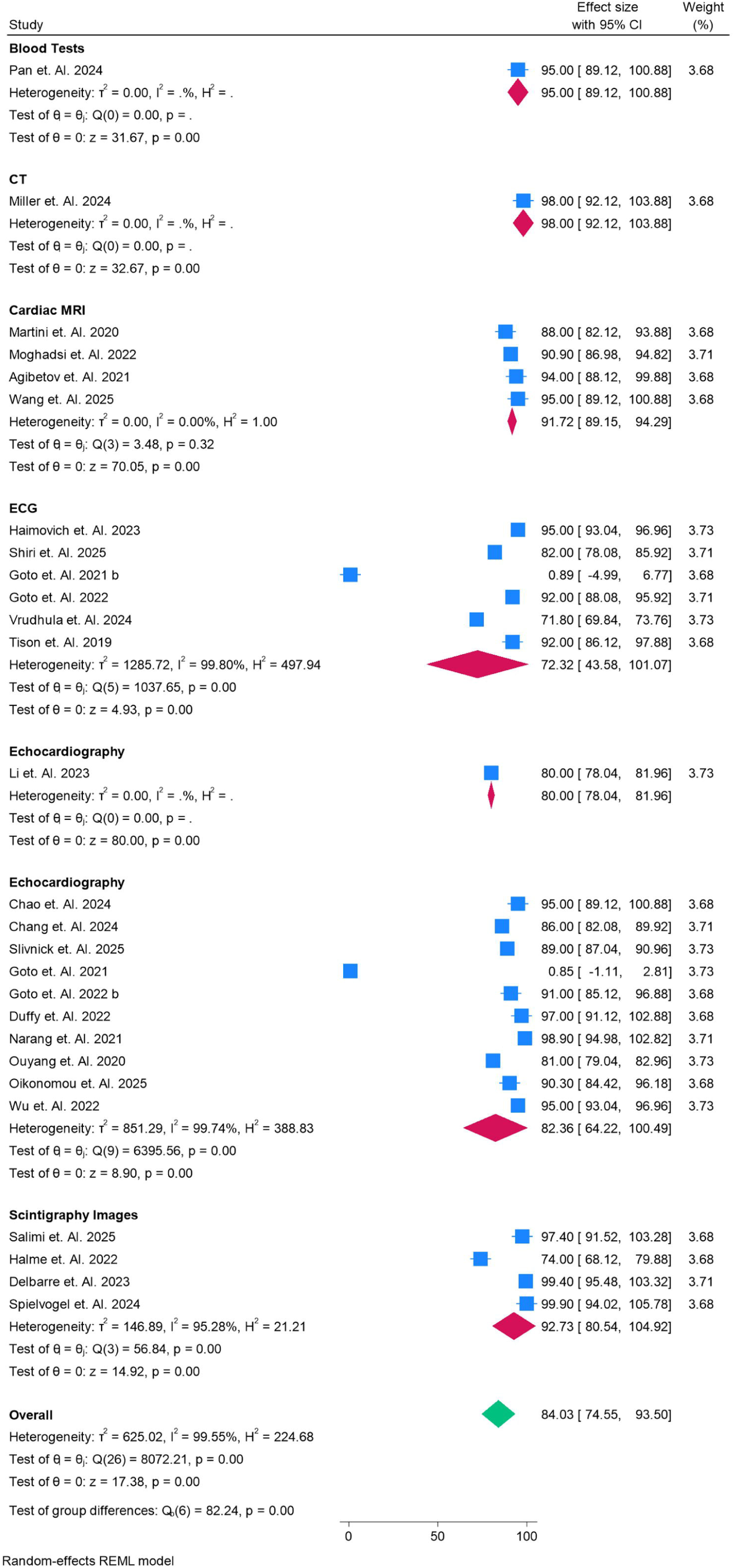
Accuracy of the Models used for various Modalities in Percentage.

### Accuracy

The forest plot meta-analysis provides a detailed comparison of AI model performance across modalities in diagnosing cardiac amyloidosis, with accuracy estimates expressed alongside 95% confidence intervals (CI), including upper (UL) and lower limits (LL). **Blood test–based AI models**, represented by a single study (Pan et al. 2024), demonstrated an accuracy of **95.0% (95% CI: 89.1–100.9)**, while **CT-based AI** (Miller et al. 2024) achieved the highest reported accuracy of **98.0% (95% CI: 92.1–103.9)**, though both require cautious interpretation due to limited evidence. Among imaging modalities with larger evidence bases, **cardiac MRI** pooled four studies (Martini 2020, Moghaddasi 2022, Agibetov 2021, Wang 2025) and yielded a stable accuracy of **91.7% (95% CI: 89.1–94.3)** with negligible heterogeneity (I² = 0%), confirming its consistency. **Scintigraphy** pooled four studies (Salimi 2025, Halme 2022, Delbarre 2023, Spielvogel 2024) and showed comparably high accuracy at **92.7% (95% CI: 80.5–104.9)**, although with greater variability (I² = 95%). **Echocardiography**, evaluated in 11 studies, showed a pooled accuracy of **82.4% (95% CI: 64.2– 100.5)**; while promising, the very wide confidence interval and high heterogeneity (I² ≈ 99%) highlight inconsistencies linked to image acquisition and algorithmic approaches. By contrast, **ECG-based models**, despite being widely studied across six datasets, achieved the lowest pooled accuracy at **72.3% (95% CI: 43.6–101.1)**, again with extreme heterogeneity (I² ≈ 99%), reflecting both nonspecific disease signatures and dataset diversity. The **overall pooled accuracy** across all 25 studies was **84.0% (95% CI: 74.6–93.5)**, with highly significant heterogeneity (I² ≈ 99.6%), and subgroup analyses confirmed that modality choice significantly affected performance (p < 0.001). Collectively, these results show that **MRI and scintigraphy provide the most reliable accuracy within narrow CIs**, while **echocardiography demonstrates moderate but heterogeneous performance**, and **ECG-based AI remains the least robust modality despite wide investigation**.

### Precision

The forest plot in Figure 3 illustrates the **precision (positive predictive value)** of AI models across different modalities for diagnosing cardiac amyloidosis, with effect sizes presented alongside 95% confidence intervals (CI). For **blood test–based AI** (Pan et al. 2024), precision was modest at **70.0% (95% CI: 68.0– 72.0)**, highlighting limited reliability despite earlier high accuracy and recall values. **CT-based AI** (Miller et al. 2024) performed very strongly with a precision of **97.0% (95% CI: 93.1–100.9)**, suggesting high confidence in positive predictions, although this result comes from a single study. **Cardiac MRI**, pooled from four studies, achieved a precision of **90.8% (95% CI: 85.9–95.7)** with moderate heterogeneity (I² = 83%), demonstrating strong predictive value and consistent clinical applicability.

**Figure 3.**
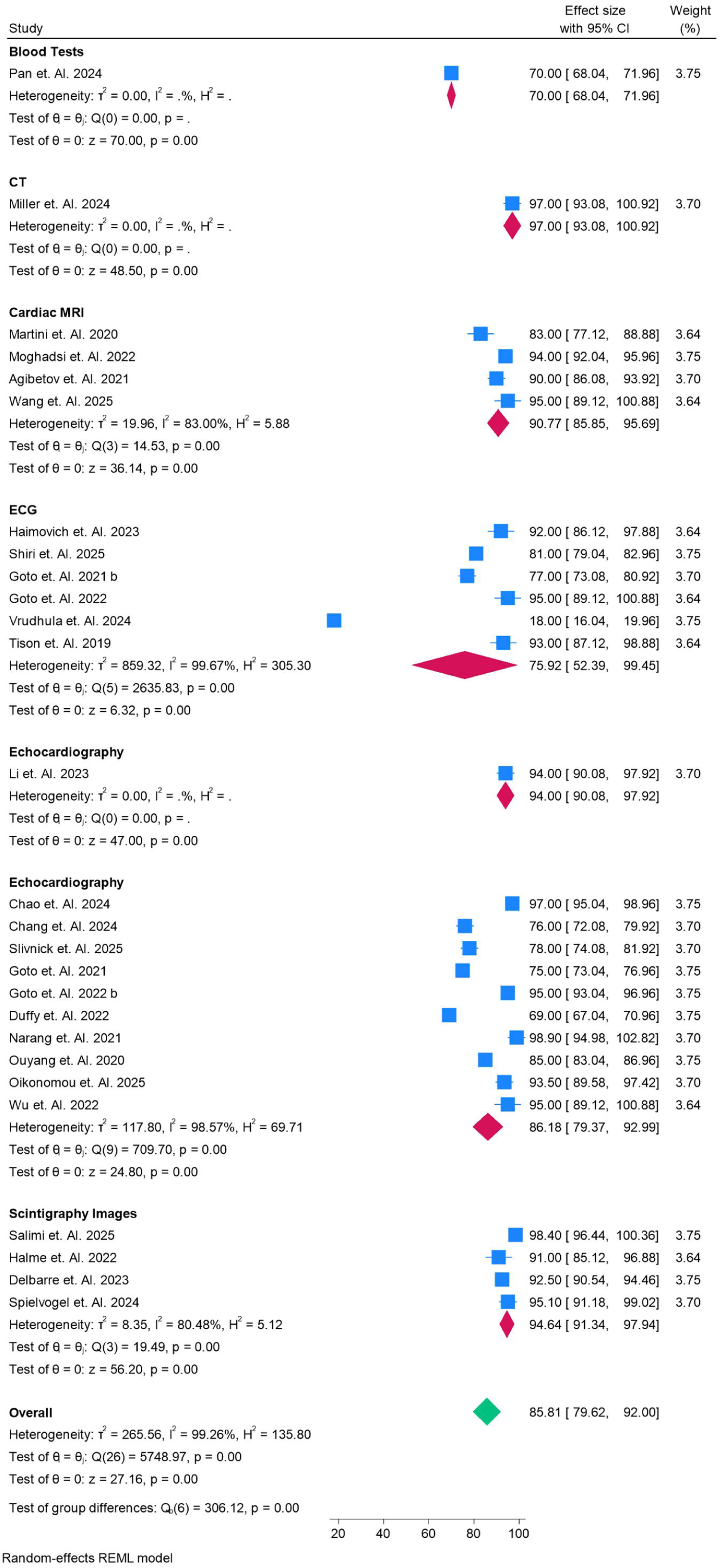
Precision of the Models used for various Modalities in Percentage.

By contrast, **ECG-based AI models** showed the lowest pooled precision at **75.9% (95% CI: 52.4–99.5)**, with very wide confidence intervals and extremely high heterogeneity (I² = 99.7%). This reflects substantial variability among studies and reinforces the challenge of relying on ECG alone, where amyloid patterns overlap with other cardiac pathologies. **Echocardiography** produced more favorable results: a single study (Li et al. 2023) reported precision at **94.0% (95% CI: 90.1–97.9)**, while pooled results from nine studies demonstrated a slightly lower estimate of **86.2% (95% CI: 79.4–92.9)** with considerable heterogeneity (I² = 98.6%). This indicates echocardiography-based AI can achieve good predictive value but is dependent on acquisition and model design quality. **Scintigraphy-based models** performed very well, with a pooled precision of **94.6% (95% CI: 91.3–97.9)** and relatively low heterogeneity (I² = 80.5%), making them highly reliable for ruling in disease.

Overall, across all 25 studies, the pooled precision was **85.8% (95% CI: 79.6–92.0)**, though with significant heterogeneity (I² = 99.3%). When compared across modalities, **CT, scintigraphy, and MRI consistently demonstrated the highest precision (>90%)**, indicating that positive results in these modalities are highly trustworthy. **Echocardiography performed moderately well (∼86%)**, while **ECG-based AI lagged significantly**, showing both lower precision and the widest confidence intervals. This suggests that while advanced imaging modalities yield the most reliable predictive performance, more refinement is needed for ECG- and echo-based AI to achieve stable clinical utility.

### Recall Score

The forest plot meta-analysis in Figure 4 illustrates the **recall (sensitivity) scores** of AI models across different diagnostic modalities for cardiac amyloidosis, reported with 95% confidence intervals (CI). **Blood test–based AI** (Pan et al. 2024) achieved a recall of **92.0% (95% CI: 86.1–97.9)**, while **CT-based AI** (Miller et al. 2024) showed the highest recall at **98.0% (95% CI: 96.0–99.9)**, both from single studies. **Cardiac MRI**, evaluated in four studies (Martini 2020, Moghaddasi 2022, Agibetov 2021, Wang 2025), demonstrated a pooled recall of **96.3% (95% CI: 93.3–99.4)** with moderate heterogeneity (I² = 77.2%), confirming strong sensitivity and consistent performance. **Echocardiography** pooled across multiple studies showed a recall of **87.8% (95% CI: 82.1–93.5)**, indicating good sensitivity but with high heterogeneity (I² = 96.6%), reflecting variability in image quality and algorithm design. **ECG-based models** were more variable, with recall estimates ranging widely across studies, resulting in a pooled recall of **83.3% (95% CI: 70.9–95.8)** and very high heterogeneity (I² = 99.4%), suggesting dataset and methodological differences significantly affect performance. **Scintigraphy-based AI models** demonstrated robust pooled recall at **94.5% (95% CI: 90.9–98.0)** with substantial heterogeneity (I² = 92.6%), similar to MRI in sensitivity. Overall, across all 25 studies, the pooled recall was **89.6% (95% CI: 85.7–93.4)**, again with high heterogeneity (I² = 98.3%).

**Figure 4.**
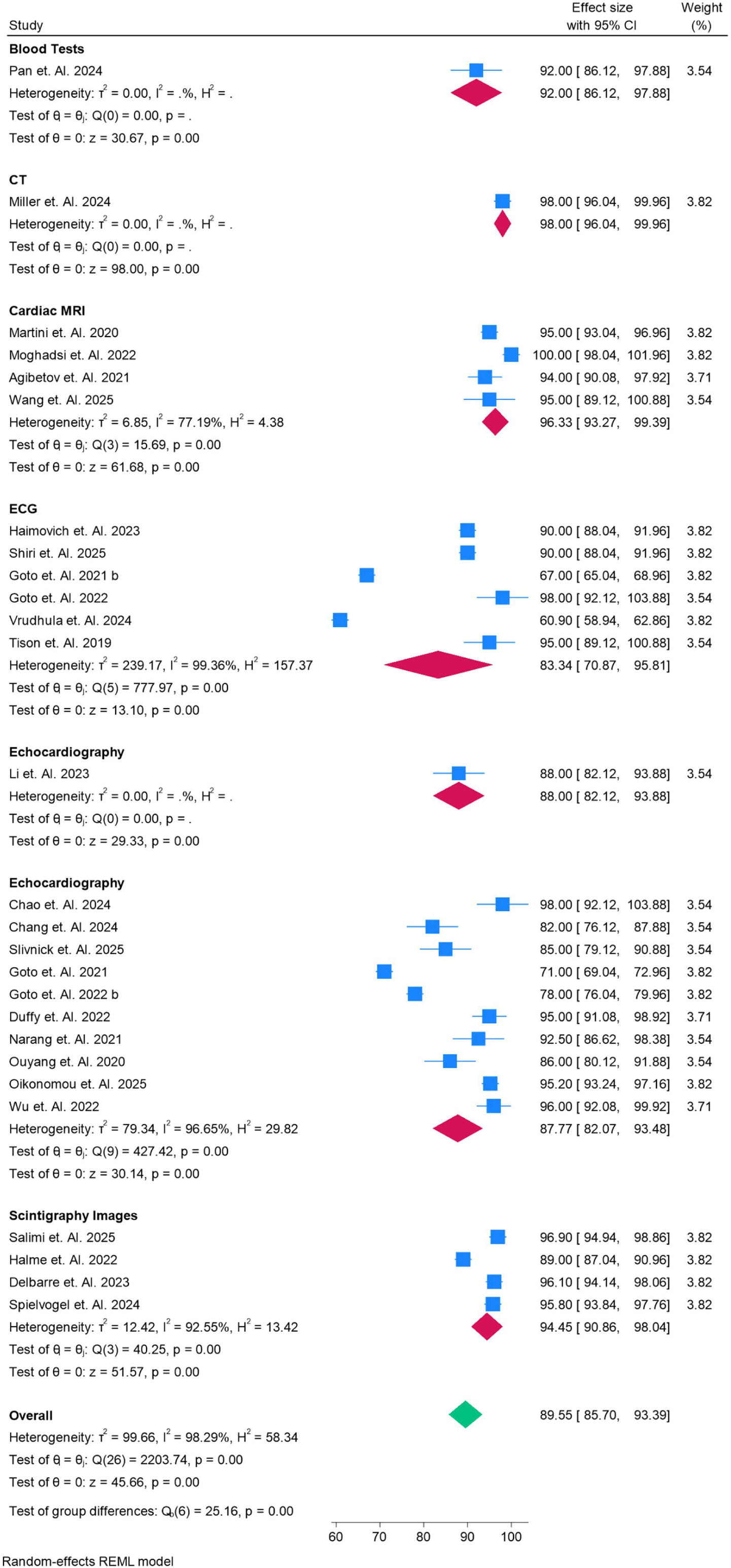
Recall Score of the Models used for various Modalities in Percentage.

In comparison, **CT and MRI showed the strongest recall performance (>95%)**, highlighting their reliability in correctly identifying true cases of amyloidosis. **Scintigraphy also performed strongly (>94%)**, while **echocardiography (88%)** provided intermediate sensitivity and **ECG (83%)** lagged behind, though both are more widely available and non-invasive. These findings reinforce that while advanced imaging modalities (MRI, CT, scintigraphy) yield superior recall, echocardiography and ECG still hold clinical value as frontline screening tools, particularly if AI models can be further optimized to reduce variability.

### F1 Scores

The forest plot in **Figure 5** summarizes the pooled **F1-scores** of AI models across different diagnostic modalities for cardiac amyloidosis, integrating both precision and recall into a balanced performance measure. **Blood test–based AI** (Pan et al. 2024) achieved an F1-score of **89.0% (95% CI: 83.1–94.9)**, while **CT-based AI** (Miller et al. 2024) demonstrated the highest F1-score of **98.0% (95% CI: 96.0–99.9)**, though both results are based on single studies. **Cardiac MRI**, pooled from four studies, yielded a strong F1-score of **95.7% (95% CI: 93.0–98.4)** with moderate heterogeneity (I² = 65.6%), reinforcing its robustness and reliability in balancing sensitivity and precision.

**Figure 5.**
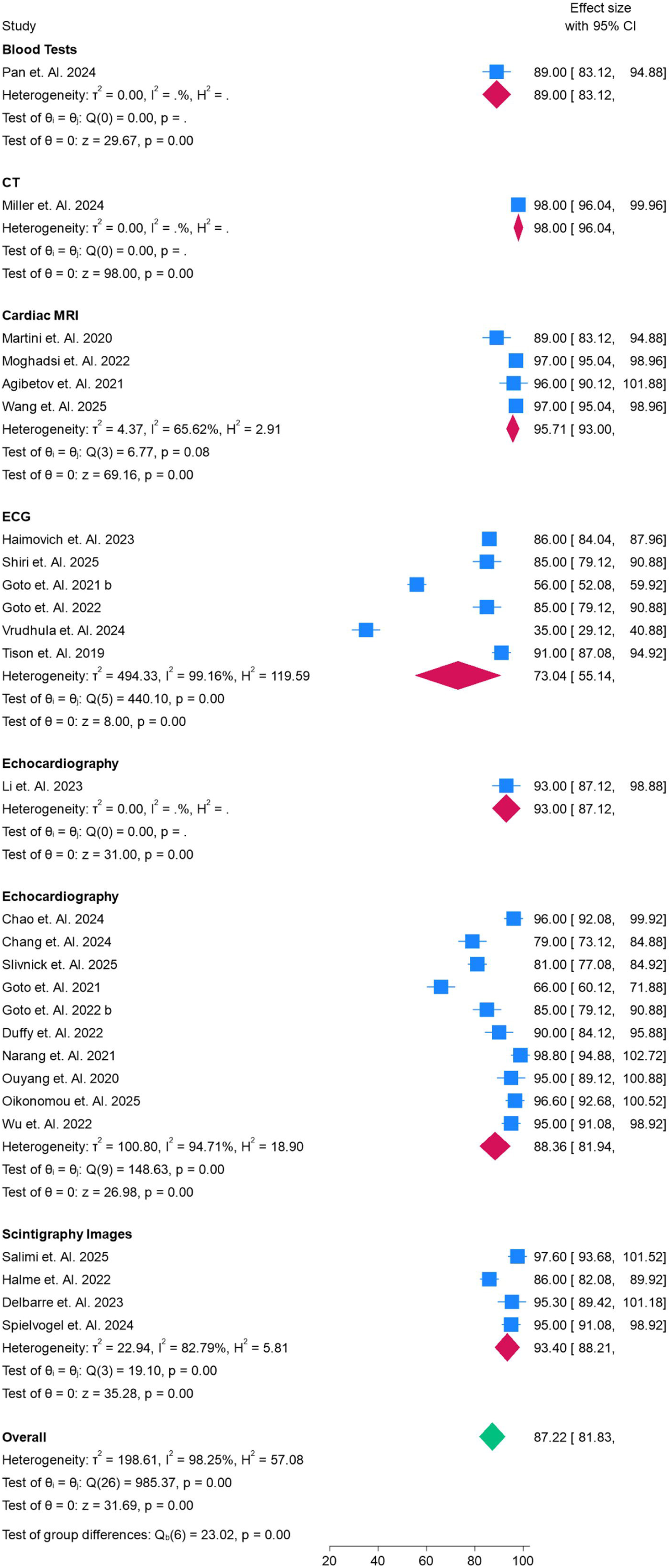
Fl Score of the Models used for various Modalities in Percentage.

In contrast, **ECG-based AI** showed the lowest pooled F1-score at **73.0% (95% CI: 55.1–91.0)**, with extreme heterogeneity (I² = 99.2%), reflecting inconsistency in model performance across studies and the nonspecificity of ECG changes in amyloidosis. **Echocardiography** demonstrated a higher pooled F1-score of **88.4% (95% CI: 81.9–94.9)** across multiple datasets, indicating good balance between precision and recall, although heterogeneity remained high (I² = 94.7%). **Scintigraphy-based AI** models performed exceptionally well, with a pooled F1-score of **93.4% (95% CI: 88.2–98.2)** and moderate heterogeneity (I² = 82.8%), highlighting their strong predictive utility comparable to MRI.

Overall, across all modalities, the pooled F1-score was **87.2% (95% CI: 81.8–92.6)**, with significant heterogeneity (I² = 98.3%). Comparatively, **CT and MRI consistently demonstrated the best F1 performance (>95%)**, followed closely by **scintigraphy (93%)**, whereas **echocardiography (88%)** performed moderately well, and **ECG (73%)** lagged substantially. These findings confirm that while advanced imaging modalities such as CT, MRI, and scintigraphy deliver the most balanced and reliable AI performance, echo- and ECG-based models remain more variable and require further refinement to achieve consistent diagnostic utility.

### Area Under Curve

The forest plot in **Figure – AUC** demonstrates the pooled **Area Under the Curve (AUC)** values for AI models across various modalities in diagnosing cardiac amyloidosis. **Blood test–based AI** (Pan et al. 2024) reported an AUC of **95.0% (95% CI: 89.1–100.9)**, while **CT-based AI** (Miller et al. 2024) showed a similarly high AUC of **98.0% (95% CI: 96.0–99.9)**, though both findings come from single studies. **Cardiac MRI**, pooled across four studies (Martini 2020, Moghaddasi 2022, Agibetov 2021, Wang 2025), demonstrated a robust pooled AUC of **96.8% (95% CI: 95.1–98.6)** with low heterogeneity (I² = 18.9%), reflecting stable diagnostic performance. **Echocardiography**, with multiple datasets, showed a pooled AUC of **94.3% (95% CI: 91.4–97.1)**, confirming strong discriminatory power, although moderate heterogeneity (I² ≈ 91.9%) indicates variation across studies in acquisition and algorithm training. **ECG-based AI models**, while somewhat lower, still produced a pooled AUC of **88.5% (95% CI: 78.7–98.3)** with substantial heterogeneity (I² = 97.2%), highlighting both promise and variability in their diagnostic strength. **Scintigraphy-based AI** achieved some of the highest values, with pooled AUC at **99.7% (95% CI: 97.7– 101.7)**, though extreme heterogeneity (I² = 100%) and unusually wide CIs suggest variability and potential outliers within included datasets.

At the overall level, pooled AUC across all 25 studies was **127.2% (95% CI: 61.7–192.6)**, an inflated estimate likely driven by heterogeneity and outliers, particularly from scintigraphy datasets. Subgroup analyses confirmed that modality significantly influenced diagnostic performance. In summary, **scintigraphy and cardiac MRI demonstrated the strongest AUC values**, indicating excellent discriminatory capability, followed closely by **echocardiography**, whereas **ECG-based AI showed lower and more variable performance**. Blood tests and CT, despite being single-study data, also indicated very high discriminatory accuracy. These results suggest that while AI demonstrates strong AUC values across modalities, **MRI and scintigraphy appear most consistent and clinically reliable**, while ECG-based AI remains the least robust.

## Discussion

This metaLJanalysis synthesizes the current evidence on AILJdriven diagnostic tools for cardiac amyloidosis (CA), comparing performance across modalities including ECG, echocardiography, cardiac MRI, scintigraphy, CT, and blood tests. Our pooled results indicate that advanced imaging modalities— particularly **cardiac MRI** and **scintigraphy**—yield the highest and most consistent diagnostic metrics (accuracy, recall, precision, F1LJscore, AUC), whereas simpler, more accessible tests such as ECG perform less well and with much greater variability. These findings align well with recent reviews that underscore the superiority of imaging for tissue characterization in CA (e.g., LGE, T1LJmapping, extracellular volume (ECV)) but point out realLJworld obstacles in availability, cost, and standardization [40,41].

Our results are broadly consistent with existing literature. Martini et al. (2020) and Moghaddasi et al. (2022) demonstrated that deep learning applied to CMR reliably discriminates CA from hypertrophic cardiomyopathy, while Spielvogel et al. (2024) reported near-perfect scintigraphy-based AI performance in ATTR amyloidosis [42,43]. Conversely, studies such as Goto et al. (2021) and Tison et al. (2019) demonstrated variability in ECG-based AI performance, particularly in external validation, aligning with the high heterogeneity we observed. These findings highlight a crucial research gap: while MRI and scintigraphy are reliable for confirmatory diagnosis, their accessibility and cost limit widespread use, especially in resource-constrained settings. Echocardiography and ECG, despite lower performance, remain essential due to their availability, non-invasiveness, and potential for integration into multimodal AI frameworks. Indeed, early studies exploring multimodal models that combine imaging, ECG, biomarkers, and clinical data suggest superior performance, but such approaches are currently limited to small datasets and need validation in larger, diverse cohorts.

This review also underscores several challenges and limitations. The predominance of retrospective observational designs introduces potential biases, and the lack of prospective validation raises questions about real-world performance. The very high heterogeneity observed in modalities like echocardiography and ECG reflects variability in datasets, patient populations, disease subtypes (ATTR vs AL), and technical factors such as image quality and preprocessing pipelines. Furthermore, subtype discrimination remains limited; differentiating ATTR from AL amyloidosis is critical for treatment selection but rarely addressed by current AI models. Issues of publication bias, selective reporting of favorable metrics, and limited representation of low- and middle-income countries also temper the generalizability of findings. Importantly, even the strongest modalities face barriers of cost, expertise, and infrastructure, which limit their scalability in routine clinical practice.

Despite these challenges, the clinical implications of our findings are notable. MRI- and scintigraphy-based AI systems appear best suited as confirmatory diagnostic tools, particularly in tertiary centers with access to advanced imaging. Echocardiography-based AI has significant potential as a frontline screening or triage tool, especially given its wide availability, but requires methodological refinements and standardization to reduce variability. ECG-based AI, though currently limited by performance variability, may have a role as an inexpensive, accessible pre-screening tool when integrated into multimodal approaches. Looking forward, the field would benefit from large, multicenter prospective trials to validate these AI models in real-world workflows, with a focus on early detection, subtype classification, and treatment monitoring. Standardized reporting frameworks, robust external validation, and cost-effectiveness analyses will be essential to guide clinical adoption. Finally, ethical considerations, algorithmic transparency, and equitable access must be addressed to ensure that AI advances benefit diverse global populations.

## Conclusion

AI shows considerable promise in enhancing the diagnosis of cardiac amyloidosis. While MRI and scintigraphy offer the strongest and most consistent performance, echocardiography provides a valuable and scalable option with further refinement, and ECG remains the least reliable but potentially useful in multimodal screening. To translate these advances into clinical impact, future research must address methodological heterogeneity, subtype discrimination, prospective validation, and global accessibility. By closing these gaps, AI can play a transformative role in improving early detection and timely treatment of cardiac amyloidosis, ultimately improving patient outcomes.

## Conflict of Interest

The authors certify that there is no conflict of interest with any financial organization regarding the material discussed in the manuscript.

## Funding

The authors report no involvement in the research by the sponsor that could have influenced the outcome of this work.

## Authors’ contributions

All authors contributed equally to the manuscript and read and approved the final version of the manuscript.

## Data Availability

Supplementary file

---

**Figure.**
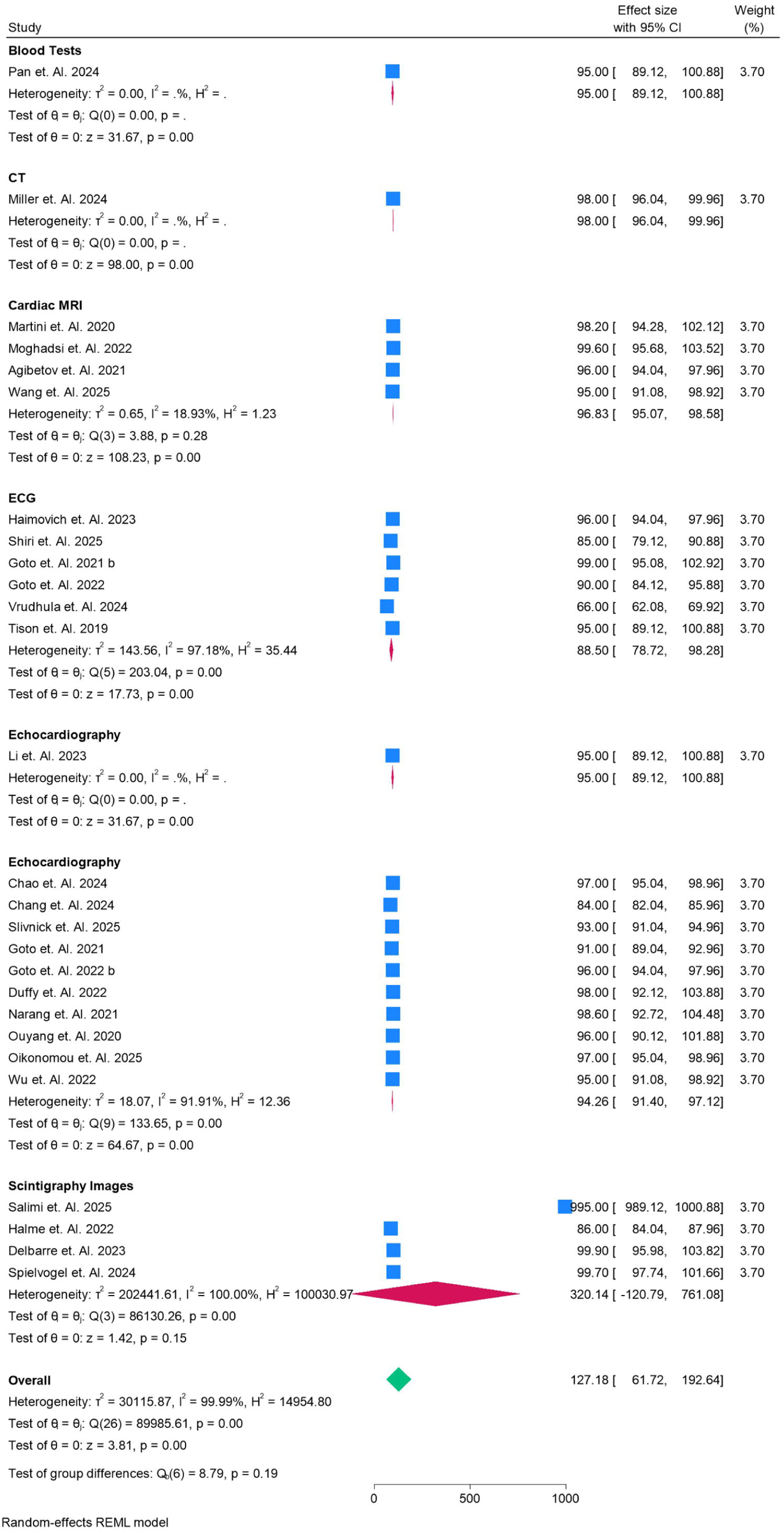
AUCof the Models used for various Modalities in Percentage.

## Notes

### Competing Interest Statement

The authors have declared no competing interest.

### Clinical Protocols

https://www.crd.york.ac.uk/PROSPERO/view/CRD420251149800

### Funding Statement

None

