## supplementary file for "Artificial Intelligence in Cardiac Amyloidosis: A Systematic Review and Meta-Analysis of Diagnostic Accuracy Across Imaging and Non-Imaging Modalities"

Supplementary Files

Search String

(("Cardiac Amyloidosis"[MeSH] OR "amyloid heart disease" OR "cardiac amyloidosis" OR "transthyretin amyloidosis" OR "light chain amyloidosis")

AND

("Artificial Intelligence"[MeSH] OR "Machine Learning"[MeSH] OR "Deep Learning" OR "Neural Network*" OR "Convolutional Neural Network" OR "CNN" OR "ResNet" OR "Transformer" OR "random forest" OR "support vector machine" OR "AI model*")

AND

("Diagnosis"[MeSH] OR "diagnostic accuracy" OR "screening" OR "classification" OR "prediction"))

AND

(observational study[pt] OR cohort[tiab] OR "case control"[tiab] OR "cross-sectional"[tiab] OR "retrospective"[tiab] OR "prospective"[tiab]))

Table S1. Data Findings and Summary of Studies

| **Ref Number** | **Author Year** | **Country** | **Total Sample** | **Male** | **Female** | **Design of Model** | **Data Origin** | **Objective** | **Type of Data** | **GRADE** | **Key Findings** | **Limitations** |
| --- | --- | --- | --- | --- | --- | --- | --- | --- | --- | --- | --- | --- |
| 13 | Martini et. Al. 2020 | Italy | 206 | 137 | 69 | CNN | **Fondazione Toscana Gabriele Monasterio, Pisa, Italy** | To evaluate a deep learning approach using CMR late gadolinium enhancement (LGE) images for diagnosing cardiac amyloidosis | Cardiac MRI | Moderate | A CNN analyzing CMR LGE images achieves **high diagnostic accuracy (AUC 0.982)** for cardiac amyloidosis, comparable to expert-level manual interpretation | Small, single-center dataset with **no external validation**, limited generalizability |
| 14 | Haimovich et. Al. 2023 | USA | 50709 | 29778 | 20931 | CNN | Mass General Brigham healthcare system | To evaluate whether AI-enabled ECG analysis can **automatically detect and classify left ventricular hypertrophy (LVH) etiologies** | ECG | Moderate | **LVH-Net AI model using ECGs accurately classifies LVH etiologies and outperforms traditional ECG rules** | data from two US hospitals, older white population, possible phenotype bias, under-recognition of rare LVH causes |
| 15 | Shiri et. Al. 2025 | Switzeerland | 263 | 149 | 114 | ML | Bern University Hospital, Switzerland | o develop and evaluate **AI-based multimodality ML models** using pre-TAVI routine data to detect **transthyretin amyloid cardiomyopathy (ATTR-CM)** in patients with severe aortic stenosis | ECG and Scintigraphy | Moderate | **CT strain-based ML models** achieved the highest diagnostic accuracy (AUC 0.85), outperforming multi-modality models for detecting ATTR-CM | **Single-center, unbalanced dataset** with limited external validation |
| 16 | Salimi et. Al. 2025 | Multicenter | 3747 | 1873 | 1874 | Segmentation and Classification | Multiple Sites | To develop and evaluate a fully automated AI pipeline for detecting and scoring transthyretin amyloid cardiomyopathy (ATTR-CM) in total-body scintigraphy images | Scintigraphy Images | Moderate | AI-based fully automated deep learning pipeline achieved excellent accuracy (AUC >0.95) for detecting and scoring ATTR-CM from scintigraphy across multi-center datasets | Highly imbalanced datasets and reliance on binary/2-class scoring instead of full 4-class Perugini grading; external dataset #6 lacked ground truth |
| 17 | Halme et. Al. 2022 | Finland | 1334 | 1089 | 245 | CNN | **Finnish nuclear medicine units** | To develop and evaluate CNN models for automated detection and classification of transthyretin amyloidosis (ATTR) from 2D bone scintigraphy images | Scintigraphy Images | Moderate | Custom CNNs (Linear and Residual) accurately detect and classify ATTR on bone scintigraphy, performing equal to or better than state-of-the-art models | Small number of positive ATTR cases and imbalanced dataset reduce generalizability |
| 18 | Chao et. Al. 2024 | USA | 381 | 278 | 103 | CNN | **Mayo Clinic Rochester** | To develop and validate a transthoracic echocardiography-based deep learning model to differentiate **constrictive pericarditis (CP)** from **cardiac amyloidosis (CA, representative of restrictive cardiomyopathy)** | Echocardiography (apical 4-chamber view, TTE) | High | A ResNet50-based deep learning model using a standard apical 4-chamber TTE view can accurately differentiate CP from CA with excellent internal AUC (0.97) and good external performance (0.84) | Retrospective design, small external dataset, potential referral bias, and limited generalization beyond CP vs CA |
| 19 | Delbarre et. Al. 2023 | France | 3048 | 1376 | 2767 | CNN | Henri Mondor University Hospital (Créteil) and Lille University Hospital | To develop and validate a CNN-based model to automatically detect significant cardiac uptake (Perugini grade ≥2) on WBS to identify patients at risk of cardiac amyloidosis | Scintigraphy Images | High | A CNN model reliably detects abnormal cardiac uptake (Perugini ≥2) on bone scintigraphy with near-perfect AUC (0.999) | The model cannot distinguish amyloidosis-related from non-amyloidosis cardiac uptake, especially Perugini grade 1 images |
| 20 | Moghadsi et. Al. 2022 | Germany | 107 | 65 | 42 | ML | Heart and Diabetes Center North-Rhine Westphalia, Ruhr-University Bochum, Germany | To assess the diagnostic performance of supervised ML algorithms using multi-chamber cardiac strain and function from CMR for detection of cardiac amyloidosis | Cardiac MRI | Moderate | Machine learning with SVM RBF kernel achieved excellent diagnostic accuracy for cardiac amyloidosis based on multi-chamber strain and function without contrast | Single-center retrospective study with small sample size and mostly advanced-stage CA patients, limiting generalizability and early-stage detection |
| 21 | Chang et. Al. 2024 | USA | 636 | 440 | 196 | RF | Cedars-Sinai Medical Center, Los Angeles | To evaluate whether a random forest model using routine echocardiographic measurements can detect cardiac amyloidosis | Echocardiography (apical 4-chamber view, TTE) | Moderate | A random forest model using 19 routine echocardiographic parameters detected cardiac amyloidosis with good accuracy (AUC 0.84) | Single-center study with high missingness for key parameters (e.g., GLS), and lack of external validation |
| 22 | Slivnick et. Al. 2025 | Multicenter | 2719 | 1524 | 1004 | CNN | Mayo Clinic Rochester | To evaluate an AI model using echocardiographic apical 4-chamber views for detecting cardiac amyloidosis (CA) | Echocardiography (apical 4-chamber view, TTE) | High | The AI model reliably differentiates CA from phenotypic mimics using a single apical 4-chamber echo view with AUC 0.93 | Retrospective design and potential bias from dataset selection; real-world prospective validation still needed |
| 23 | Goto et. Al. 2021 | USA | 480 | 240 | 240 | CNN | Brigham and Women’s Hospital (training), validated at Massachusetts General Hospital, UCSF, Northwestern, and Keio University | To develop and validate AI models using ECG and echocardiography for early, automated detection of cardiac amyloidosis | Electrocardiograms (ECG) and echocardiograms (Echo) | High | AI-based ECG and Echo models can detect cardiac amyloidosis with high accuracy and outperform expert cardiologists | Possible undiagnosed cases in control groups may have introduced mislabeling bias |
| 24 |  |  |  |  |  |  |  |  |  | High |  |  |
| 25 | Goto et. Al. 2022 | Multicenter | 62814 | 31407 | 31407 | CNN | Massachusetts General Hospital, University of California San Francisco, Brigham and Women’s Hospital (US), and Keio University Hospital (Japan) | To develop federated learning–based CNN models using ECG and echocardiogram data to detect hypertrophic cardiomyopathy (HCM) with robust generalizability across institutions | **ECG and Echocardiogram** (not Cardiac MRI or Scintigraphy) | Moderate | Federated learning significantly improved the generalizability of ECG and echocardiogram CNN models for HCM detection compared with single-institution training | Potential inclusion of undiagnosed HCM in controls and reliance on tertiary-care data limit generalizabilit |
| 26 |  |  |  |  |  |  |  |  |  | Moderate |  |  |
| 27 | Duffy et. Al. 2022 | USA | 23745 | 16360 | 7385 | ResNet3D | **Stanford Health Care**, **Cedars-Sinai Medical Center**, and **Unity Imaging Collaborative** | To evaluate whether a deep learning algorithm can automatically quantify left ventricular hypertrophy and predict its underlying causes (e.g., HCM, cardiac amyloidosis). | **Echocardiography videos** (parasternal long-axis and apical 4-chamber views) | High | The deep learning algorithm outperformed human experts in automated echocardiographic measurement and accurately distinguished causes of LV hypertrophy. | The model was trained on curated tertiary-care clinic cohorts, limiting generalizability across more diverse populations |
| 28 | Narang et. Al. 2021 | USA | 240 | 139 | 101 | CNN | **Northwestern Memorial Hospital** and **Minneapolis Heart Institute** | To test whether novices could obtain diagnostic-quality echocardiograms using AI-guided deep-learning software | **Echocardiography (Echo, TTE – transthoracic echocardiography)** | Moderate | AI guidance enabled novices to acquire echocardiograms of diagnostic quality in >90–98% of patients | Small sample size, limited to 2 academic centers, and lack of a true novice control group without AI |
| 29 | Li et. Al. 2023 | USA | 586 | 339 | 247 | Deep Learning | Mayo Clinic Arizona | To develop and validate an **automated deep learning echocardiography framework** to classify etiologies of increased left ventricular wall thickness (HCM, CA, HTN/others) | Echocardiography | Moderate | A **fusion deep learning model using six echo views** accurately differentiates hypertrophic cardiomyopathy, cardiac amyloidosis, and hypertensive heart disease. | Single-center retrospective dataset without external validation, limiting generalizability |
| 30 | Miller et. Al. 2024 | Canada | 299 | 190 | 109 | Deep Learning | University of Calgary | To evaluate a **deep learning approach for fully automated quantification of 99mTc-pyrophosphate SPECT/CT in cardiac amyloidosis** | CT | Moderate | **Deep learning–based automated quantification of 99mTc-pyrophosphate SPECT/CT achieves near-perfect diagnostic accuracy for cardiac amyloidosis and predicts adverse outcomes** | **Limited by single time-point imaging (3h only), no widespread biopsy confirmation, and reliance on correct CT/SPECT registration** |
| 31 | Agibetov et. Al. 2021 | Austria | 502 | 240 | 262 | CNN | Vienna General Hospital, Medical University of Vienna | To test whether CNNs applied to cardiac MRI can **automatically detect cardiac amyloidosis** and improve diagnostic accuracy | Cardiac MRI | Moderate | Fine-tuned CNN models on CMR images achieved high diagnostic performance (ROC AUC up to 0.96), comparable to human experts in diagnosing cardiac amyloidosis | Single-center design with limited generalizability; imbalance in patient sex distribution; small number of amyloidosis cases; failure to capture early/preclinical disease |
| 32 | Vrudhula et. Al. 2024 | USA | 341989 | 239392 | 102597 | CNN | Cedars-Sinai Medical Center, Los Angeles, California, USA | To evaluate how case and control selection impacts AI-based ECG waveform models for screening cardiac amyloidosis | **Electrocardiogram (ECG)** | Moderate | AI models trained on less curated ICD-based amyloid cases performed comparably to models trained on highly curated clinic cases in population-level screening | Lack of external validation, reliance on ICD codes (possible misclassification), and potential undiagnosed amyloidosis in controls |
| 33 | Ouyang et. Al. 2020 | USA | 10030 | 4885 | 5145 | CNN | Stanford Health Care, Cedars-Sinai Medical Center, and Unity Imaging Collaborative | To develop and validate a **video-based deep learning model (EchoNet-Dynamic) for beat-to-beat left ventricular ejection fraction (EF) assessment from echocardiogram videos** | Echocardiography | High | EchoNet-Dynamic **outperformed human experts in EF estimation, with higher reproducibility and generalizability across hospitals** | Model performance may vary with **variable image quality and non-cardiologist acquisitions; external validation was limited to U.S. academic centers** |
| 34 | Oikonomou et. Al. 2025 | USA | 33127 | 17393 | 19229 | 3D-ResNet18 | Yale–New Haven Health System | To develop and validate AI models using POCUS videos to detect under-diagnosed cardiomyopathies (hypertrophic and transthyretin amyloid cardiomyopathy). | Echocardiology | High | AI-POCUS can detect hypertrophic and transthyretin amyloid cardiomyopathies years before clinical diagnosis, with prognostic implications. | Retrospective design, under-diagnosis leading to possible false positives, POCUS not directly used for model training |
| 35 | Spielvogel et. Al. 2024 | Multicenter | 16241 | 6748 | 9493 | CNN | Medical University of Vienna, Austria | To **develop and validate an AI system for automated detection of cardiac amyloidosis-suggestive uptake in scintigraphy and assess its prognostic value** | Scintigraphy Images | High | AI-based screening reliably detects cardiac amyloidosis on scintigraphy, outperforming physicians, and provides independent prognostic value for mortality | **Lack of histology/SPECT-CT confirmation for all cases and limited consecutiveness in some cohorts (PYP, HMDP)** |
| 36 | Pan et. Al. 2024 | China | 261 | 194 | 67 | ML | Wuhan Asia Heart Hospital, Wuhan, Hubei Province, China | To develop and validate a machine learning model using routine blood tests to improve early detection of cardiac amyloidosis in patients with LVH | Blood Tests | Moderate | An XGBoost-based ML model using routine blood tests accurately detects cardiac amyloidosis with AUC 0.95, outperforming traditional biomarkers | The study had imbalanced CA vs control samples, possible undiagnosed CA cases in controls, no external validation, and loss of data due to exclusion of tests with >60% missing values |
| 37 | Tison et. Al. 2019 | USA | 36186 | 18093 | 18093 | CNN | UCSF MUSE ECG database | To develop an automated, interpretable ECG profiling framework for disease detection, tracking, and discovery | ECG | Moderate | Machine learning-based ECG profiling can accurately detect and track cardiac structure and diseases with high interpretability | Data was derived from a **single medical center** and limited to **normal sinus rhythm ECGs**, restricting generalizability |
| 38 | Wang et. Al. 2025 | USA | 89 | 42 | 47 | Res-CRNN | **Washington University in St. Louis** | To evaluate whether AI-assisted compressed sensing CINE (AI-CS-CINE) accelerates cardiac MRI acquisition and improves workflow compared to conventional CINE | Cardiac MRI | Moderate | AI-CS-CINE reduced CMR scan/reconstruction time by 84% while maintaining excellent diagnostic agreement with conventional cine MRI | Limited generalizability – single-center study on one 1.5T scanner, predominantly cardio-oncology and healthy volunteers |
| 39 | Wu et. Al. 2022 | China | 138 | 87 | 51 | ML | **The First Affiliated Hospital, Zhejiang University School of Medicine, Hangzhou, China**. | To develop machine learning models using echocardiography and speckle-tracking echocardiography to differentiate **cardiac amyloidosis (CA)** from **hypertrophic cardiomyopathy (HCM)**. | Echocardiography | Moderate | Machine learning models using echocardiography and 2D-STE achieved **excellent accuracy (AUC > 0.9)** in differentiating cardiac amyloidosis from hypertrophic cardiomyopathy. | Single-center, small-sample study with limited generalizability and only echocardiographic data considered. |

Risk of Bias using QUADAS 2.0

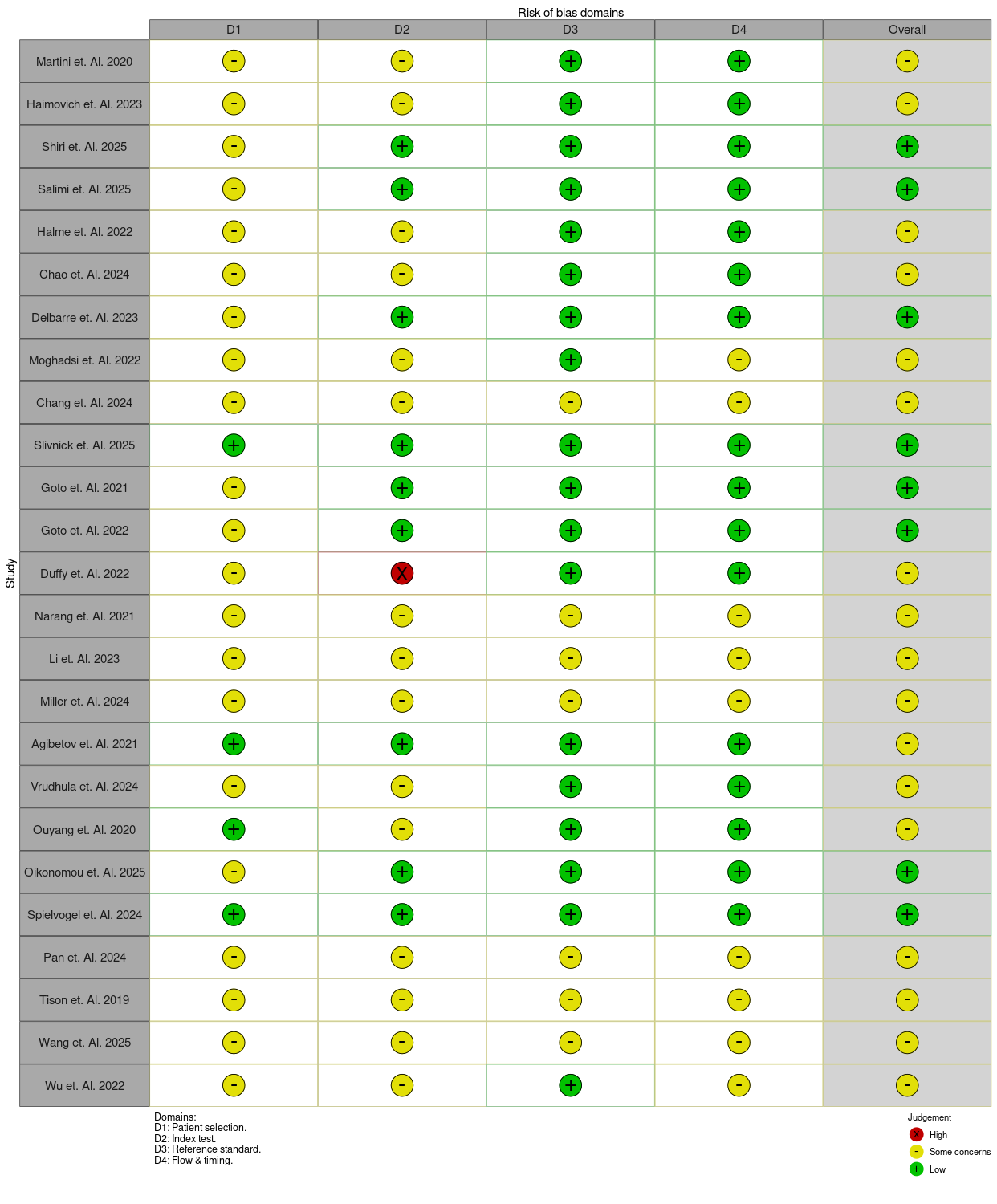
